# Type construction in qualitative health research and its relevance in complementary and integrative medicine. Two studies about patients’ experiences with herbal medicine preparations

**DOI:** 10.1101/2022.06.13.22276352

**Authors:** Claudia Canella, Balz Wolfensberger, Claudia M. Witt

## Abstract

In this article, we explore the method of type construction in qualitative health research in the field of complementary and integrative medicine. We applied type construction to research questions in phytotherapy about the everyday life experiences of patients using two specific herbal medicine preparations. In total, 21 patients participated in two consecutive qualitative studies. The collected data incorporated patient diaries, face-to-face interviews and pre- and posttreatment questionnaires. A type construction approach was applied for data analysis. In the Passiflora-study, three distinct biographical narrative types attributed to different experiences when using a specific ethanolic extract of *Passiflora incarnata* were identified. In the Angelica-study, four types of thematic dimensions of experiences with a specific ethanolic extract of *Angelica archangelica* were identified. Type construction in qualitative health research can contribute to evidence-based phytotherapy and complementary and integrative medicine in general by supporting shared decision making and individualized treatment approaches.

## Introduction

Qualitative content analysis is rooted and widely used in the German speaking countries and has started to be discussed internationally for some years now (1, 2). Type construction is an approach within qualitative content analysis aiming at building multidimensional ideal types from the data (3, 4). As experiences, preferences, needs and values of patients are one of the three foundations of evidence-based medicine (cochrane.de; 5), identifying individual and collective dimensions of stakeholders’ experiences is crucial in qualitative health research (4, 6, 7). Therefore, type construction seems to be an especially suitable approach towards this goal, but there is still a respective research gap in the field of phytotherapy and complementary and integrative medicine (CIM) in general (1, 3, 4).

### Methodology of type construction

Firstly, the overall process of type construction often starts with a content analysis of the collected qualitative data (3, 4). Secondly, individual cases are reconstructed and compared with each other. Finally, multidimensional ideal types are built (4, 8). The range of individual cases can include the description of single events in life, for example the experiences of the use of herbal medicine in our studies, to the narrative of a whole biography (4, 8). The aim of case comparison is to identify dimensions of collective experiences towards the research question(s) (4, 8). A typical process of type construction is not linear but iterative, where the single steps are repeatedly performed in relation to answer the research question(s). Figure 1 visualizes such a typical process which is described according to Kuckartz in the following (8). Usually, the process starts with defining the relevant dimensions towards type construction based on the collected data. The data are often structured by performing a coding process. Based on this analysis, types are constructed, and the individual cases are attributed to the types. Changing to a meta-level, a typology is inductively constructed, also considering context factors which inform the typology.

**Figure 1:**
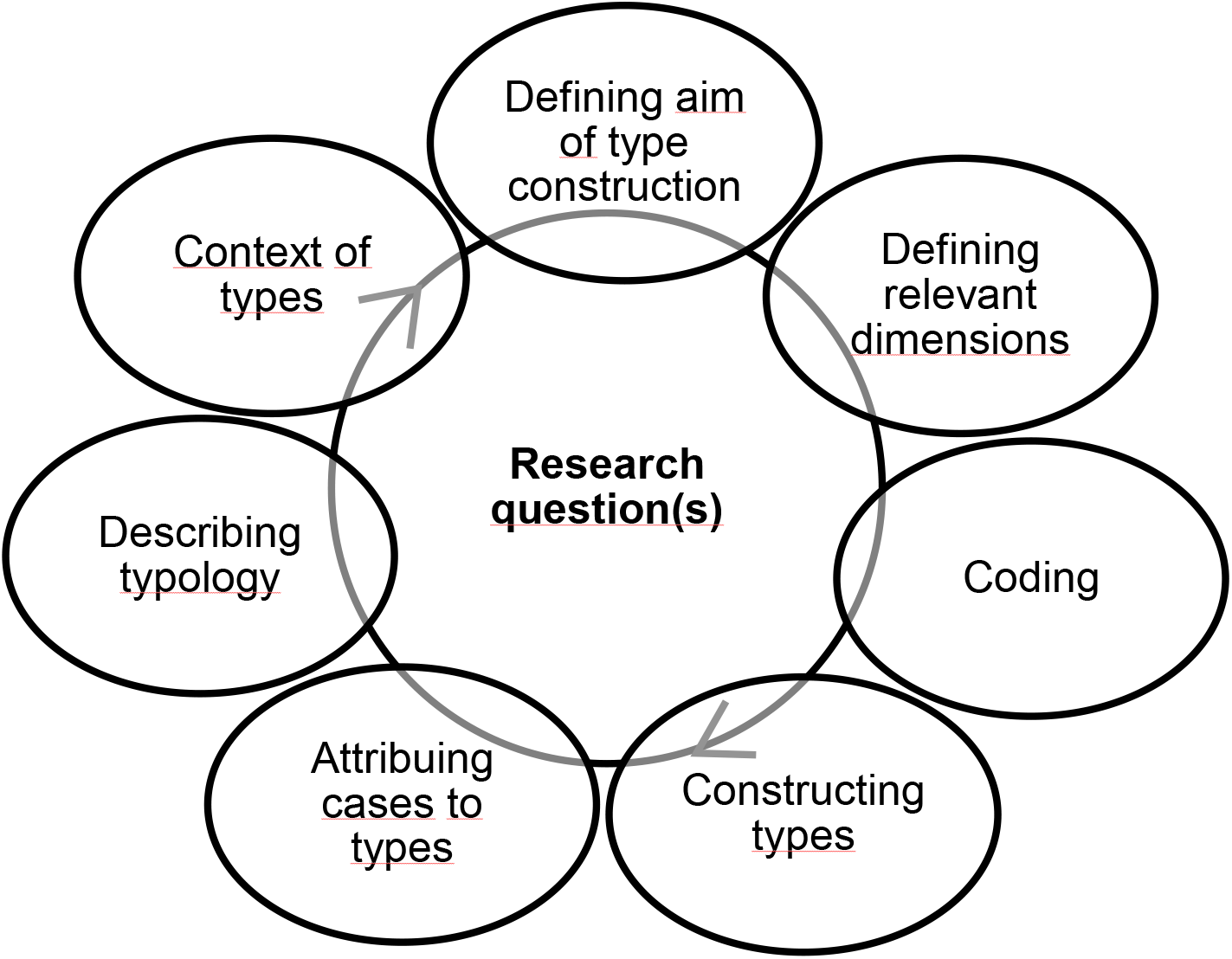
Typical process of type building in qualitative content analysis according to Kuckartz (8/,153)

### Two qualitative studies about Passiflora incarnata and Angelica archangelica

In two consecutive qualitative studies, we explored how patients experience the use of two herbal medicine preparations, *Passiflora incarnata* and *Angelica archangelica*, and which expectations, values and views the patients relate to their use of them. *Passiflora incarnata* is frequently prescribed for anxiety, sleep disorders and restlessness in phytotherapy (9, 10). *Angelica archangelica* is frequently prescribed for gastro-intestinal complaints, often stress-induced, exhaustion and as expectorant in phytotherapy (http://www.theplantlist.org, last accessed February 12th, 2021; 11). For both studies, we used specific ethanolic herbal extracts produced by the Swiss manufacturer Ceres Heilmittel AG (Swissmedic-nr.: 400827 *Passiflora incarnata*, 400830 *Angelica archangelica*, production according to HAB § 3a and PhEur; 12). The usual dosage recommendation for the two researched ethanolic herbal extracts are three times three drops per day.

### Aim

In this article, we aim at exploring the analytic method of type construction in qualitative health research in the field of complementary and integrative medicine (CIM). We share our insights from applying type construction to research questions in phytotherapy about patients’ experiences attributed to the use of two herbal medicine preparations, *Angelica archangelica* and *Passiflora incarnata*. We will discuss the results by comparing them to existing descriptions of the two medicinal plants. Finally, we will reflect on the relevance of type construction within the context of phytotherapy and CIM in general.

## Methods

Both studies were explorative and observational in nature and were consecutively conducted in the German speaking part of Switzerland. The Passiflora-study was conducted between 2015 to mid-2017 and the Angelica-study between mid-2017 to 2019. Both studies did not fall under the regulation of the Human Research Act of Switzerland which was confirmed by the Ethics Committee of Zurich, Switzerland. We obtained written informed consent for participation and scientific publication from all the participants (for ethics details see separate sections at the end of the article).

The two studies only slightly differed in their approach to data collection and analysis. The proceeding of the first study about *Passiflora incarnata* is already published in detail in the Journal of Ethnopharmacology (13). Therefore, in this article, we concentrate on reporting the overall approach to both studies and details from the second study on *Angelica archangelica* that we adapted according to the learnings from the Passiflora study. In this article, we focus on the patients’ data and on reporting the type construction process, but we did also perform a participant observation during the production process of the ethanolic herbal extracts in question and asked the referring medical doctors to fill in a questionnaire about their prescription routine and dosage recommendations of the two herbal extracts in their usual care setting.

### Qualitative Data Collection

Medical doctors specialized in phytotherapy referred patients to the study coordinator (first author) after having prescribed *Passiflora incarnata* or *Angelica archangelica* manufactured by Ceres Heilmittel AG for the first time to individual patients during their usual care setting. The consultation, the prescription and the follow ups between the medical doctors and the patients were independent of the studies. In the next step, the study coordinator informed the patients about the study and included them consistent with the inclusion and exclusion criteria (see table 1) and after the patients provided informed consent.

**Table 1:** Inclusion and exclusion criteria

| Inclusion criteria | Exclusion criteria |
| --- | --- |
| Intake of *Passiflora incarnata* or *Angelica archangelica* produced by Ceres Heilmittel AG after prescription from a physician specialized in phytotherapy | Persons who are unable to follow the data collection process due to limited German language skills |
| Patients > age 18 | Persons who are directly linked to Ceres Heilmittel AG |
| Written informed consent | Persons who already used *Passiflora incarnata* or *Angelica archangelica* produced by Ceres Heilmittel AG within the past 5 years |

Table 2 visualizes the process of data collection. The patients filled in a questionnaire within the first two weeks after their first intake of the herbal extract. They answered questions about their socio-demographics, their medical history, symptoms that lead to the prescription of the herbal medicine preparation in question and about their expectations towards the intake of the ethanolic herbal extract. In the third week after their first intake, patients started to write a diary at least once a week about their daily routines, experiences, thoughts, and feelings with the intake of the herbal medicine preparation for three weeks in a row.

**Table 2:**
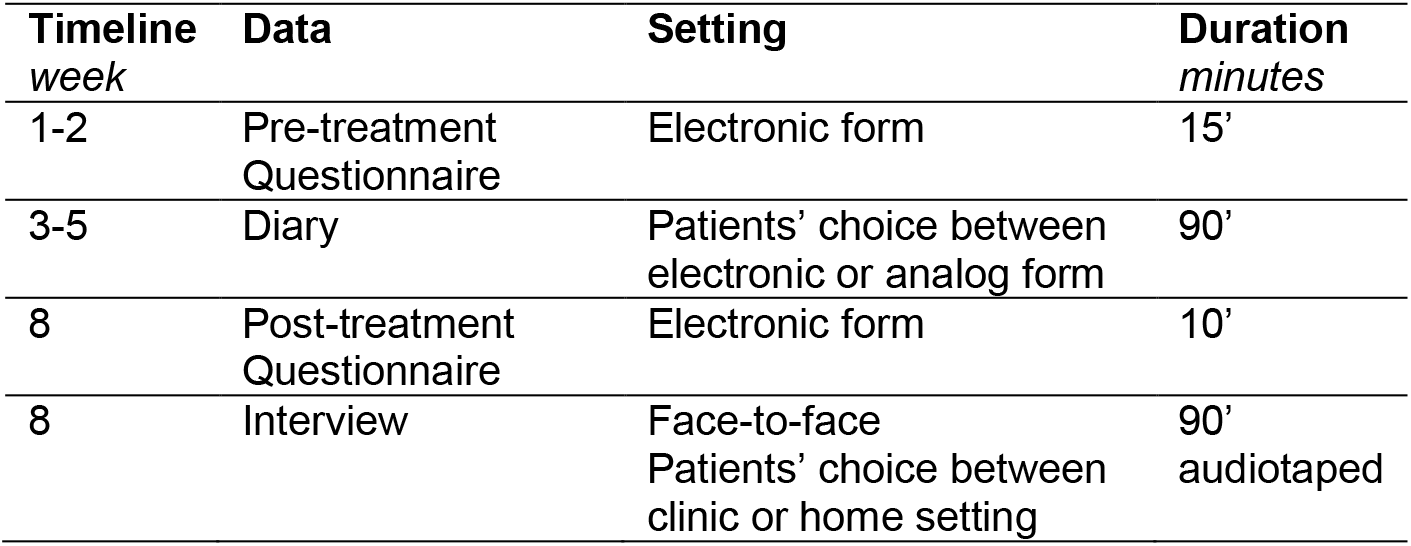
Data collection process, timeline, setting and duration

| Timeline<br>week | Data | Setting | Duration<br>minutes |
| --- | --- | --- | --- |
| 1-2 | Pre-treatment<br>Questionnaire | Electronic form | 15' |
| 3-5 | Diary | Patients' choice between<br>electronic or analog form | 90' |
| 8 | Post-treatment<br>Questionnaire | Electronic form | 10' |
| 8 | Interview | Face-to-face<br>Patients' choice between<br>clinic or home setting | 90'<br>audiotaped |

After eight weeks, the patients completed a questionnaire about the development of their symptoms, and to what extent their expectations towards the use of the herbal medicine preparation have been met. In the same week, the study coordinator/researcher (first author) conducted a concluding face to face semi-structured interview with the patients about their overall experiences with the use of the ethanolic herbal extract in question.

We continued with purposeful sampling (14) until we reached a saturated spectrum of expressed topics and experiences. We also aimed at covering an expectable diversity in socio-demographics and medical history of the patients within the resources of the study (14). In the Passiflora-study, there was one non-participation without declaration of a reason and one dropout due to a private issue which was not related to the study. This dropout was not included in the analysis. In the Angelica-study, there was no dropout, but four non-participations, two without indicating a reason, one due to lack of time and one declared not having enough energy to participate.

### Qualitative Data Analysis

We used MAXQDA software (Release 11.1.2 for the Passiflora-study and 18.2.4 for the Angelica-study) for qualitative data analysis. In both studies, the questionnaires were analyzed by descriptive statistics and all the interviews were transcribed verbatim. Whereas the diary and interview data in the Passiflora-study required analytic methods towards reconstructing biographical narrative types, the data sets in the Angelica-study pointed more towards reconstructing types of thematic dimensions of experiences. That is why the approach to analysis slightly differed in the two studies. The detailed proceeding in the Passiflora-study has already been published (13). For the Angelica-study, according to the process of type building described above (4, 8), we performed a thematic coding of of the interview transcripts, where the coding units were units of meaning, and codes were built inductively from the data and using within- and across-case comparison (4, 15). Based on the codings, a summary for every case was written. Subsequently, we aggregated across-case types of experiences, considering all the data and context information (4, 8).

For every step in the analysis, we performed an intersubjective validation process between two researchers (first and second author) to work towards reliability and robustness of the data analysis (14, 16). Step by step, one researcher (first author) performed the analysis and the other (second author) validated it. After every step, the two researchers met to discuss the analysis until a consensus was reached. If necessary, the analysis was revised according to the validation discussion. Intersubjective validation (first, second and third author) was also performed while conceptualizing data collection. Throughout the whole process, we practiced memo writing about our thoughts, feelings, and assumptions to minimize interpreting the data with implicit values and hypotheses. Finally, we discussed the findings with all the researchers and clinicians of our Institute. The participants did not comment on the transcripts but were informed on the findings.

## Results

### Participants

In total, 21 patients, twenty women, one man, with an age range from 24 to 84 participated in the two studies. Table 3 summarizes the participants’ characteristics. All patients suffered from comorbidities and were using co-medication and different conventional and CIM treatments during the studies.

**Table 3:**
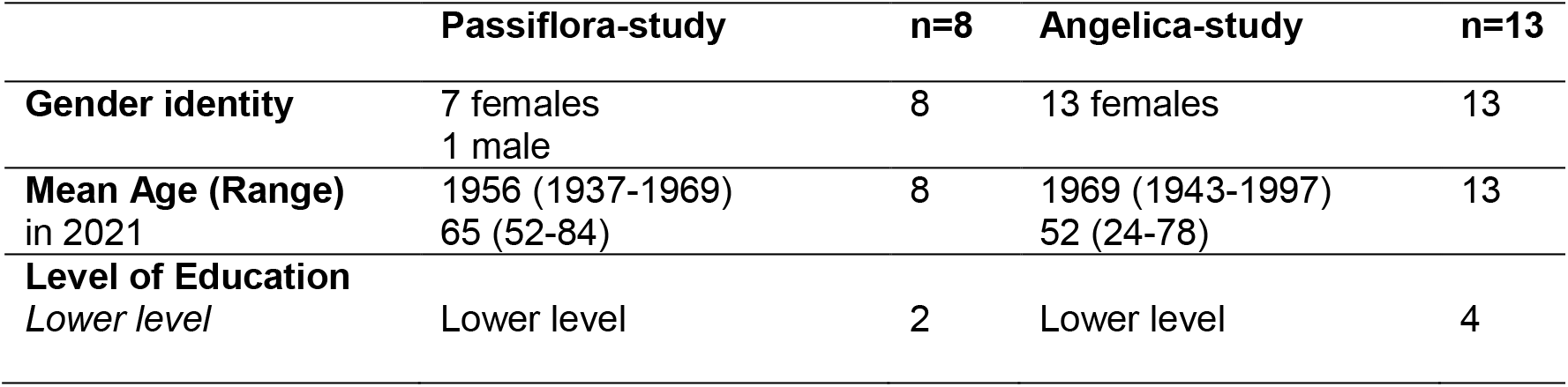
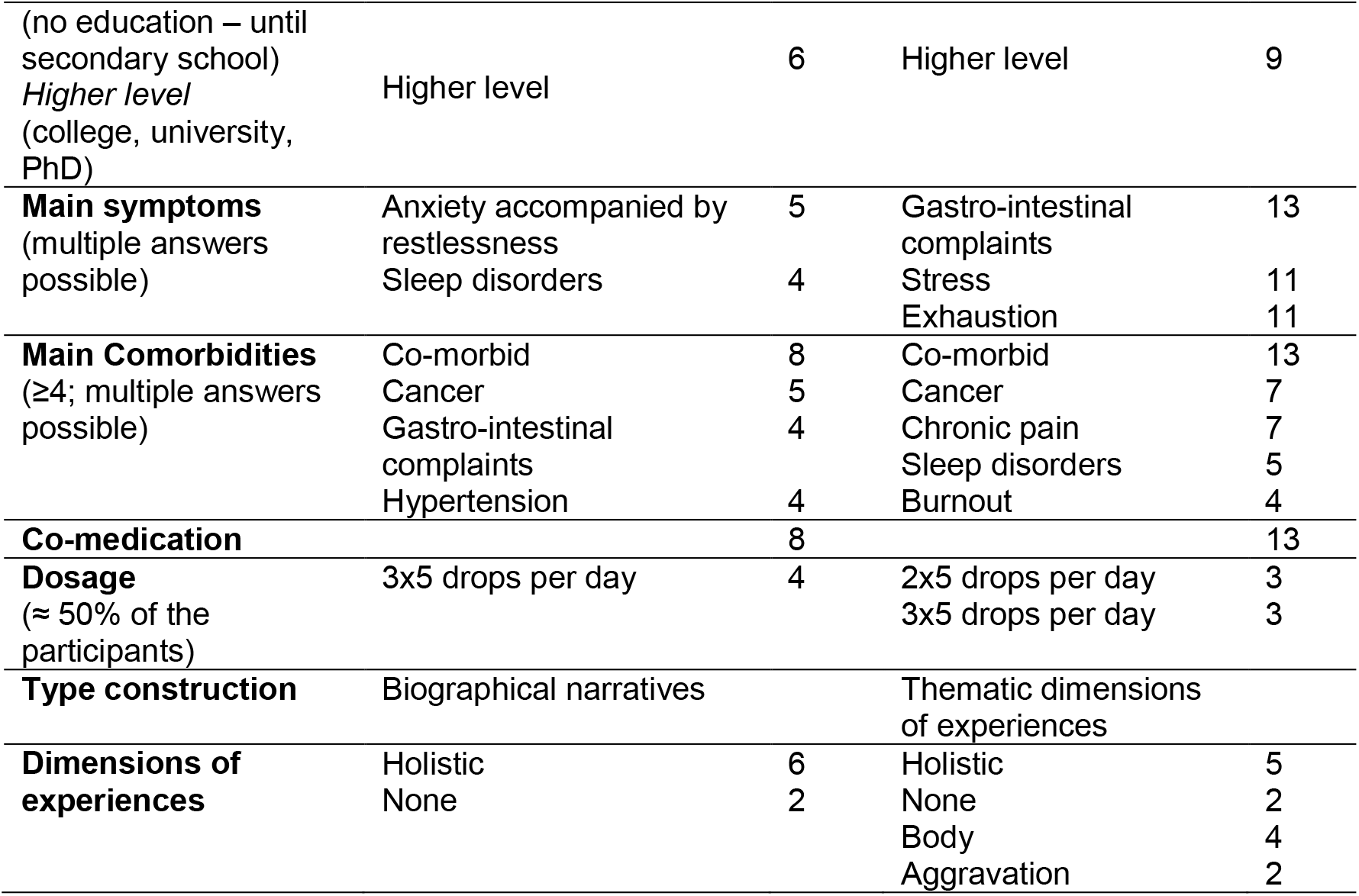
Participants’ characteristics – Comparison between the Passiflora-study and the Angelica-study

The main symptoms that led to the prescription of one of the two ethanolic herbal extracts in question were for the Passiflora-study, anxiety accompanied by restlessness and sleep disorders, and for the Angelica-study, gastro-intestinal complaints, stress, and exhaustion. About half of the participants reported using a dosage of 2×5 drops per day or 3×5 drops per day. The other half used different dosages ranging from 1×1 drop per day to 3×8 drops per day. The applied dosages were self-reported by the patients and could differ from the prescription of their medical doctors.

### Thematic Codes of the Angelica-study

In the Angelica-study, we identified thirteen different thematic codes from the data. The codes were inductively built from the patients’ self-reported experiences of the use of the *Angelica archangelica* extract. Thereby, the reported experiences can be both, negative or positive in the view of the patients. For most of the codes, we used the patients’ own denominations of their experiences, for example, several patients labelled some of their experiences as “spiritual”. Table 4 provides an overview over the codes, their definitions, and a representative quote.

**Table 4:** Overview over thematic codes, code definitions and representative quotes from participants of the Angelica-study

| Thematic Code | Code definition | Representative quote |
| --- | --- | --- |
| Physiological | Perceived physiological symptoms, changes, and processes in general. | “Now, I know that I immediately get a quiet stomach. Although I wanted to use the herbal drops more for my mind, it is the body that reacts” (KL, diary day 3). |
| Esthesia | Organoleptic experiences: taste, smell, optics, haptic, acoustics. | “[It tastes]...slightly bitter, not unpleasant...like a plant root” (RK, interview, 130). |
| Mind | Experiences of one’s own mind. | “It seems like a psychological effect of the drops is manifesting itself, a strengthening effect.” (KW, diary day 1). |
| Emotional | Emotional experiences and perceptions. | “It [the stomach] is simply calm and not painful anymore...I feel safe” (KL, interview, 552). |
| Spiritual | Spiritual experiences. | “I feel accompanied...They [the Angelica drops] have something strong...a strong personality, something that is strengthening me...it strengthens me to get inside me and having my feet on the ground” (MU, interview, 8, 10). |
| Personal development | Descriptions of one’s own personality and how it changed over time and while using Angelica. | “I feel strengthened and courageous, also in showing myself publicly. I don’t have to hide or being ashamed...I feel strong...With great certainty I can say: that was a growth” (KW, interview, 86, 100, 104). |
| Narratives | Personal narratives related to one’s own medical condition: causes, explanations, hypothesis, interpretations, and developments. | “I have a lot of narratives...for example, when I get gastro-intestinal complaints, I say: that is because I did not have enough time in the morning to relax...” (JI, interview, 441, 447). |
| Everyday actions | Descriptions of noticed changes in the own everyday actions. | “Through repeating the experience changes...Rituals are helpful to me...a predefined time, in a certain manner, always the drops in the glass. It is like an exercise in mindfulness” (AS, interview, 83, 85, 91). |
| Expectations | Expectations towards the use of Angelica and its expected effects. Descriptions of how the expectations changed over time. | “[I expect] relaxation in the gut area” (AS, diary day 1 & 2). |
| Evaluation | Explicit self-reported evaluation of perceived negative or positive effects of the use of Angelica. | "After the intake it burns on my tongue, it is very unpleasant...I wake up the next day or in the night and my tongue is so dry...and swollen" (LM, interview, 425, 427). |
| Intake and Dosage | Self-declarations of intake and dosage. | "Instead of three times a day, I sometimes took it only two times a day" (VA, interview, 30). |
| Information | Indications of searched or received information about the herbal extract or the producer. | "I entered it to the internet and had a look at how the plant looks like" (VA, interview, 476). |
| CIM | Statements about CIM in general. | "I am the kind of person who says that I first try it with an herbal product or with homeopathy or with anthroposophical remedies, and when it does not work out, I take a pill" (DF, interview, 16). |

The thematic codes range from the levels on which the experiences are perceived, i.e. in the body or in the mind, to interpretations of the own personality, i.e. personal development, and reports about everyday actions. In addition, expectations and evaluations are represented as codes as well as indications of intake and dosage and received information about the herbal extracts or the producer.

### Type construction in the Angelica-study – Description of four types

In the next step of the analysis, we wrote a case summary for each participant along the codes. From there, we performed a within- and across-case comparison aiming at identifying the relevant dimensions of experiences pertaining to the use of *Angelica archangelica*, individually as well as collectively. We identified six essential dimensions of experiencing the use of the *Angelica archangelica* herbal extract: body, mind, emotions, spirituality, personal development, and everyday actions. We then allocated the single cases to these six dimensions (see Table 5).

**Table 5:** Allocation of single cases to six essential dimensions of experience of the *Angelica archangelica* herbal extract. Participants’ names are pseudonyms.

| **Dimensions of experiences** | **Body** | **Mind** | **Emotions** | **Spirituality** | **Personal development** | **Everyday actions** |
| --- | --- | --- | --- | --- | --- | --- |
| Valentina Amrein | x | - | - | - | - | - |
| Melanie Berger | x | (x) | - | - | - | - |
| Jasmin Isler | - | - | - | - | - | - |
| Dorit Frenkel | - | - | - | - | - | - |
| Elsbeth Gerber | x | - | - | (x) | (x) | - |
| Renate Koch | Agg. | - | - | - | - | - |
| Silke Lehmann | x | x | x | x | x | x |
| Katrin Leu | x | x | x | x | x | x |
| Laura Moser | (x) | - | - | - | - | - |
| Antonia Schneider | Agg. | - | - | - | - | x |
| Margarete Unger | x | x | x | x | x | x |
| Katja Winter | x | x | x | x | x | x |
| Manuela Zollinger | x | x | x | - | x | x |

The symbol “x” in the table indicates that the participant attributed explicitly some of their own experiences of the use of Angelica to the respective dimension. Katja Winter for example clearly relates to a physical experience in the interview saying: “I feel a regulating aspect [of Angelica] in my stomach, it is more physical” (KW, interview, 253). The symbol “(x)” indicates that the experience needed further interpretation by the researchers to be able to attribute it to a respective dimension. Laura Moser for example began her interview with reporting that Angelica helped her with digestion: “…Concerning the digestion, I think it helped…But I also got prescribed other things…well, I also take psyllium” (LM, interview, 13-16). In the course of the interview, she further explained that she suffers less from constipation since taking Angelica, but still had cramps and because of that, for the first time, she was able to relate the constipation to her digestion and the cramps to her monthly cycle (LM, interview, 54-58). In the end, we interpreted that she had physical experiences after her relating the constipation to digestion and evaluating that her digestion improved after using Angelica.

The symbol “-” stands for no reported experiences with the use of Angelica towards the dimension in question of the participants. The acronym “Agg.” means that the participant experienced an aggravation of their complaints which they explicitly related to the use of the *Angelica archangelica* herbal extract. Renate Koch for example reported:

> “…it felt like a stone lying in my stomach…and it became hard and bulky like a huge clump pressing all over…It did not end and I decided to stop with the remedy after a week, and then it slowly, slowly got better.” (RK, interview, 67-69).

In the final step, we formed four types by identifying collective patterns of experiences with the intake of the *Angelica archangelica* extract among the participants. We noticed that the participants either reported a holistic experience including at least five or all six dimensions or only having physical experiences; either with positive associations or reporting an aggravation of their physical complaints. Two participants had no experiences which they explicitly related to the use of the Angelica herbal extract. Table 6 provides an overview over the allocation of the cases to the four types.

**Table 6:** Overview allocation of the single cases to four types of experiencing the use of *Angelica archangelica* herbal extract.

|  | **Types** | Holistic | Body | None | Aggravation | N |
| --- | --- | --- | --- | --- | --- | --- |
| **Participants (Pseudonyms)** |  |  |  |  |  |  |
| Silke Lehmann |  | x |  |  |  |  |
| Katrin Leu |  | x |  |  |  |  |
| Margarete Unger |  | x |  |  |  |  |
| Katja Winter |  | x |  |  |  |  |
| Manuela Zollinger |  | x |  |  |  |  |
| Valentina Amrein |  |  | x |  |  |  |
| Melanie Berger |  |  | x |  |  |  |
| Elsbeth Gerber |  |  | x |  |  |  |
| Laura Moser |  |  | x |  |  |  |
| Jasmin Isler |  |  |  | x |  |  |
| Dorit Frenkel |  |  |  | x |  |  |
| Renate Koch |  |  |  |  | x |  |
| Antonia Schneider |  |  |  |  | x |  |
|  |  | 5 | 4 | 2 | 2 | n=13 |

Four patients of the “holistic” type reported experiences of at least five or all six dimensions. As an example, table 7 lists representative quotes of Katja Winter who was assigned to the holistic type.

**Table 7:** Example of representative quotes of the holistic type – Katja Winter

| **Example of representative quotes of the holistic type – Katja Winter (pseudonym)** |  |
| --- | --- |
| **Dimensions of experiences** |  |
| *Body* | «I take it [Angelica drops] and then it warms down towards the belly/ it calms the belly, puts it in order. I had the impression it is regulating after stress, fast eating, a little twitching in the stomach and then it [Angelica] puts order into it.” (KW, interview, 124) |
| *Mind* | “I notice that the plant seems to put in order everything and providing a feeling of being kept together inside. Almost something like that it provides guidance” (KW, diary day 1). |
| *Emotions* | I feel very, very much strengthened and much safer...I feel emotionally calmed down.” (KW, interview, 80, 504) |
| *Spirituality* | I had a subtle feeling of being accompanied...by a plant creature...you know, ‘*Angelica archangelica*’? I thought, I might be accompanied by an angel...(laughs)...the Angelica-angel or something like it (laughs)...It had something motherly and nourishing because I experienced it as very benevolent...and at the same time demanding.” (KW, interview, 233, 235) |
| *Personal development* | “I felt completely released...I could fully immerse myself with a total inner conviction and strength. And I remember being surprised how easy and coherent that was for me.” (KW, interview, 54) |
| *Everyday actions* | It brought me to focus on sensing more precisely in different areas/ (...) what do I really feel there, you know?!” (KW, interview, 190) |

Furthermore, the participants of the holistic type often summed up their experiences to a whole rounded experience. Manuela Zollinger’s summary, for example, sounded like this:

> “…It started with Angelica…I sleep better and therefore I can recover better. I regained strength. My skin cleared up…A lot of the inner, the mind is processing in the belly…and Angelica released my digestion, silenced my stomach and gut, and made the energy flow…I feel a bit love of life now. It comes up from the stomach…the gut feeling says: ‘it is good’…I feel lighter in many ways.” (MZ, interview, 245)

They all also reflected on the role Angelica played in their process. Silke Lehmann, for example, phrased it that way:

> “I think, a rudimentary path was already there, in everything that I’m now describing to you related to my changed experiences. But sometimes you need a catalyzer to be able to implement the things the way you want them. That is the right term; Angelica is a catalyzer to me.” (SL, interview, 542-545).

Manuela Zollinger did not report spiritual experiences, but we allocated her to the holistic type nevertheless, because she described different experiences covering all other five dimensions.

The four patients of the “body” type reported distinct physical experiences with the use of *Angelica archangelica*. An example is the description of Valentina Amrein:

> “Before, the problem with flatulence was massive…I also had problems with defecation, constipation…and then I started to work more and more…and it [Angelica] really helped very well with the flatulence…if you sit in the cable car or at work and having a flatulence problem, that is very uncomfortable…You always have to be cautious not to release wind…you are a little bit under pressure then. And with the drops [Angelica] this was not a problem anymore (laughs) and I was relaxed.” (Valentina Amrein, 7-24)

We allocated Elsbeth Gerber to the “body” type although she adressed the dimensions “spirituality” and “personal development”, but she related her experiences with these dimensions more to her cancer journey than to her use of Angelica. Similarly, we proceeded with the allocation of Melanie Berger to the “body” type as she related her experiences in the dimension of the “mind” more to her recovery of a burnout before using Angelica.

There were two special cases concerning the dimension of “everyday action”. Establishing a ritual around the intake would have been very important to Jasmin Isler, but she did not achieve to establish one with the *Angelica archangelica* herbal extract (JI, interview, 212-220). Practicing a ritual around the intake was also very important to Antonia Schneider and made her feel safe. She enjoyed having a ritual with the intake of the *Angelica archangelica* herbal extract, despite experiencing an aggravation of her complaints (AS, interview, 49-51).

### Type construction in the Passiflora-study – Summary of three types

The proceeding of type construction and the detailed description of the single types in the Passiflora-study were already published elsewhere (13). Figure 2 provides an overview over the three types of the Passiflora-study. Different than in the Angelica-study, the participants of the Passiflora-study embedded their experiences with the use of the herbal extract into a biographical narrative (13).

**Figure 2:**
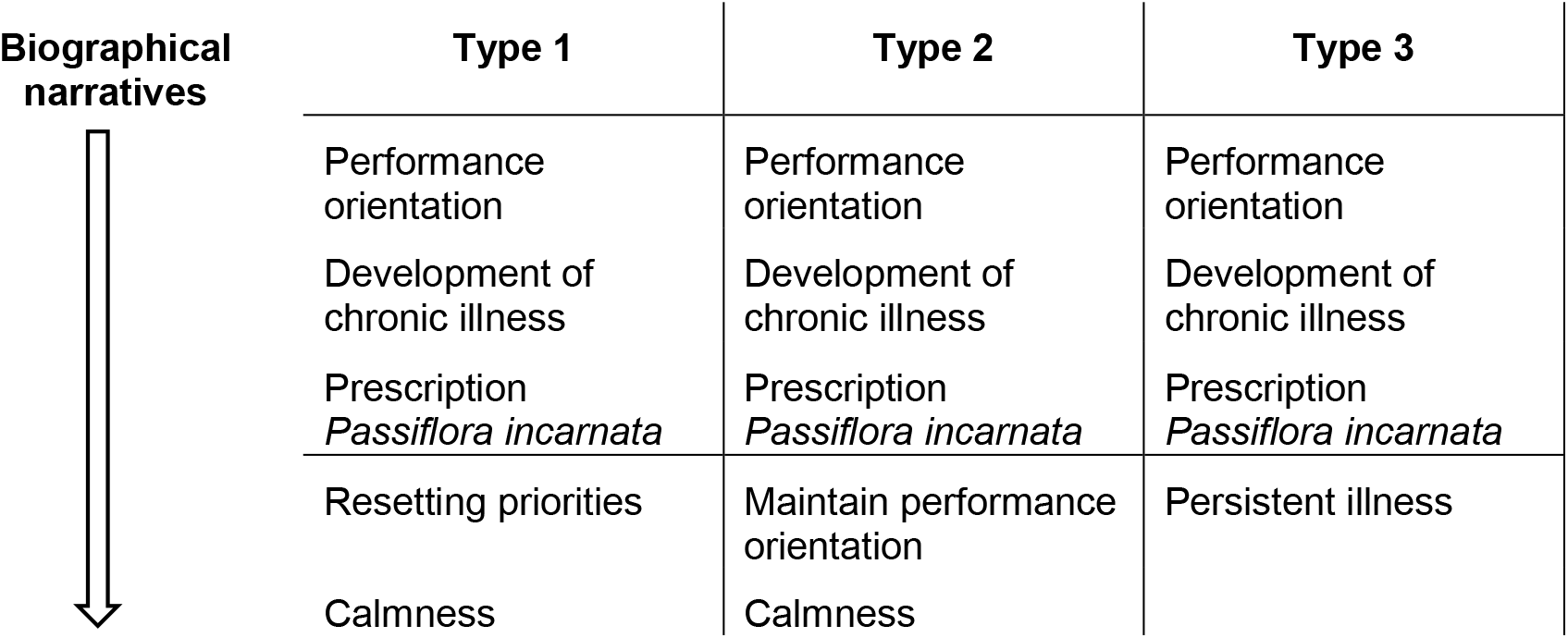
Types of the Passiflora-study (13).

All types reported that they were highly performance oriented and developed chronic illness over time (13). After the prescription of Passiflora the participants’ journeys parted. Type one patients reported a resetting of their priorities in life and attaining calmness, whereas type two patients also reported becoming much calmer while still maintaining their performance orientation (13). Like the Angelica-study, there were also participants (type three) with no experiences related to the use of the *Passiflora incarnata* herbal extract (13).

## Discussion

To discuss the relevance of type construction in phytotherapy and CIM in general, we would like to compare the findings of the two studies with already existing descriptions of the two herbal medicine plants in question. Most of the common and widely used descriptions of herbal medicine plants (e.g. 9) entail the botanical description and occurrence of the plant, its traditional use, the analysis of its phytochemistry, pharmacological activities and effects, and its indications. The descriptions of indications range from physical symptoms to emotional states and challenges of the mind. If we look at specific existing descriptions of *Passiflora incarnata* (9, 17) and *Angelica archangelica* (11, 12, 17) (see tables 8 and 9), which cover two poles of a broad range of existing descriptions, we noticed that they describe a momentum in the state of the potential patient.

**Table 8:** Overview description of the entry of indications of *Passiflora incarnata* in ESCOP (9), the description of the type 1 biographical narrative of the Passiflora-study and the description of the essence of *Passiflora incarnata* from the producer (17).

|  | **ESCOP entry indications *Passiflora incarnata*** | **Type 1 narrative of the Passiflora-Study** | **The Essence of *Passiflora incarnata*** |
| --- | --- | --- | --- |
|  | "Tenseness, restlessness, and irritability with difficulty in falling asleep" (9) | Performance orientation<br>Development of chronic illness<br>Prescription *Passiflora incarnata*<br>Resetting priorities<br>Calmness | Attaining peace of heart<br>Reconnecting with oneself<br>Aligning with the inner self<br>Becoming aware of the inner tone |
| **Representative matching quotes of the Passiflora-study** | I had always a "full brain" and was constantly "over-excited". I "processed my work while sleeping", which resulted in sleep disorders. (13) | I developed from being "a workaholic" to "selfcare", "feeling valuable" and "feeling calm". (13) | "Passiflora, I feel, is providing me with a better foundation for these changes that have come about or that have started. I feel that I am on different ground here, that my new steps on this different ground are somehow more solid. That is, I am not perpetually racking my brain anymore. Therefore, I am not that unstable any more...in everything. That is what I identify with Passiflora." (13) |

**Table 9:**
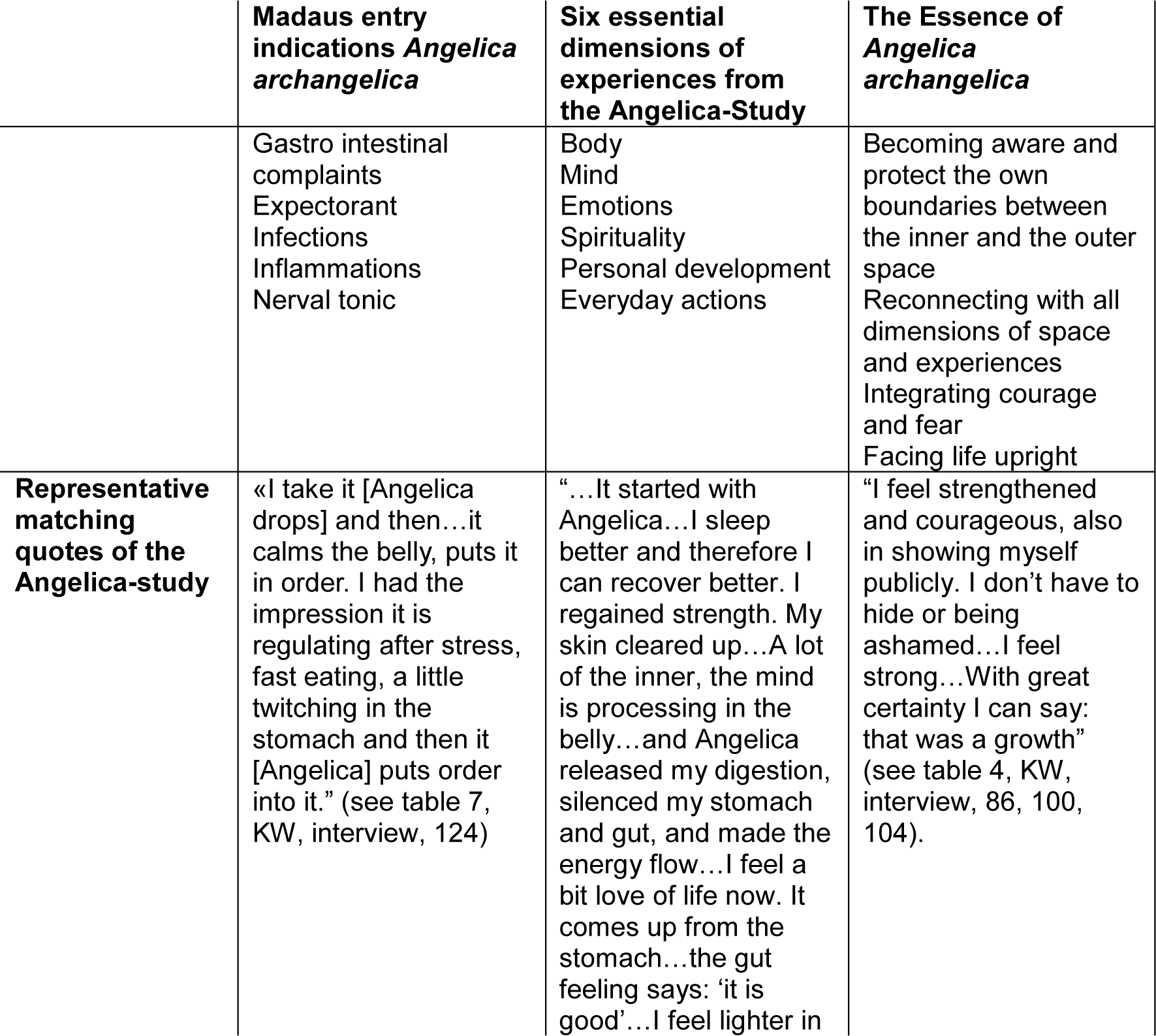
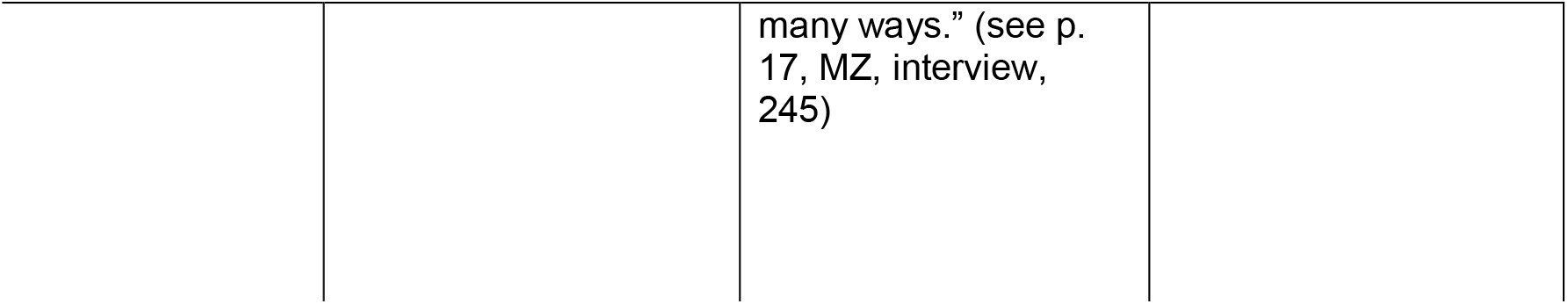
Overview description of the entry of indications of *Angelica archangelica* in Madaus (11) the description of the six essential dimensions of experiences from the Angelica-study and the description of the Essence of Angelica archangelica from the producer (17).

|  | **Madaus entry indications *Angelica archangelica*** | **Six essential dimensions of experiences from the Angelica-Study** | **The Essence of *Angelica archangelica*** |
| --- | --- | --- | --- |
|  | Gastro intestinal complaints<br>Expectorant<br>Infections<br>Inflammations<br>Nerval tonic | Body<br>Mind<br>Emotions<br>Spirituality<br>Personal development<br>Everyday actions | Becoming aware and protect the own boundaries between the inner and the outer space<br>Reconnecting with all dimensions of space and experiences<br>Integrating courage and fear<br>Facing life upright |
| **Representative matching quotes of the Angelica-study** | «I take it [Angelica drops] and then...it calms the belly, puts it in order. I had the impression it is regulating after stress, fast eating, a little twitching in the stomach and then it [Angelica] puts order into it." (see table 7, KW, interview, 124) | "...It started with Angelica...I sleep better and therefore I can recover better. I regained strength. My skin cleared up...A lot of the inner, the mind is processing in the belly...and Angelica released my digestion, silenced my stomach and gut, and made the energy flow...I feel a bit love of life now. It comes up from the stomach...the gut feeling says: 'it is good'...I feel lighter in many ways.” (see p. 17, MZ, interview, 245) | "I feel strengthened and courageous, also in showing myself publicly. I don't have to hide or being ashamed...I feel strong...With great certainty I can say: that was a growth" (see table 4, KW, interview, 86, 100, 104). |

The ESCOP (9) and the Madaus entry (11) focus on listing complaints which can potentially be treated with the herbal medicine in question, whereas the descriptions of the producer of our researched preparations (17) focus more on the healthy mind and its state after being potentially treated with the herbal medicine in question. Our type descriptions in the two studies display the process from suffering from complaints to experiencing changes on different levels after the use of the herbal medicine in question. The method of type descriptions as used here have the potential to connect the different herbal medicine descriptions. In the Angelica-study, for example, we found many reports of gastro-intestinal complaints, which are listed as the main indications in the respective entries (e.g. 11), and how they improved with the use of *Angelica archangelica*. Within the “holistic” type, we equally identified descriptions that remind of the essence of *Angelica archangelica* described by the producer of the herbal preparation used for the study (17). For example, Katja Winter who reported feeling courageous (see quote in table 4, KW, interview, 86) after using Angelica, or Margarete Unger who described feeling accompanied and grounded by the drops (see quote in table 4, MU, interview, 8, 10). The same is true for the type 1 biographical narratives of the Passiflora-study.

Being able to display the process while using the herbal medicine preparations by describing types might contribute to the clinical process in phytotherapy and CIM in general.

### Potentials and limitations of type construction in the clinical context of phytotherapy and CIM (see Figure 3)

Being aware of type descriptions can support therapeutic decision making when the medical doctor must decide which herbal medicine preparation to prescribe among the different possible suitable medical plants for the presented complaints of the patients (18, 19). For example, if patients present themselves with stress-induced gastro-intestinal complaints and the medical doctor specialized in phytotherapy ends up with a therapeutic decision to be made between several possible bitterns after having performed detailed case taking and diagnostics, having in mind the characteristics of the *Angelica archangelica* type descriptions might be helpful in this regard as a starting point to conversations with patients aiming at shared decision making (18, 19). The type descriptions could be used as a mirror where treating doctors and their patients can reflect together on the specific situation of the patient and discuss about the patients’ preferences, needs and values (20, 21). Such discussion about the patients’ preferences, needs and values would put into practice what is defined as one of the three foundations of evidence-based medicine (cochrane.de; 5). In addition, the goal of an individualized treatment approach could be strengthened by shared decision making supported by type descriptions as explained above (18, 19, 21). This is especially relevant in the field of complementary and integrative medicine as there are often different options which are equally valuable to treat the patients.

**Figure 3:**
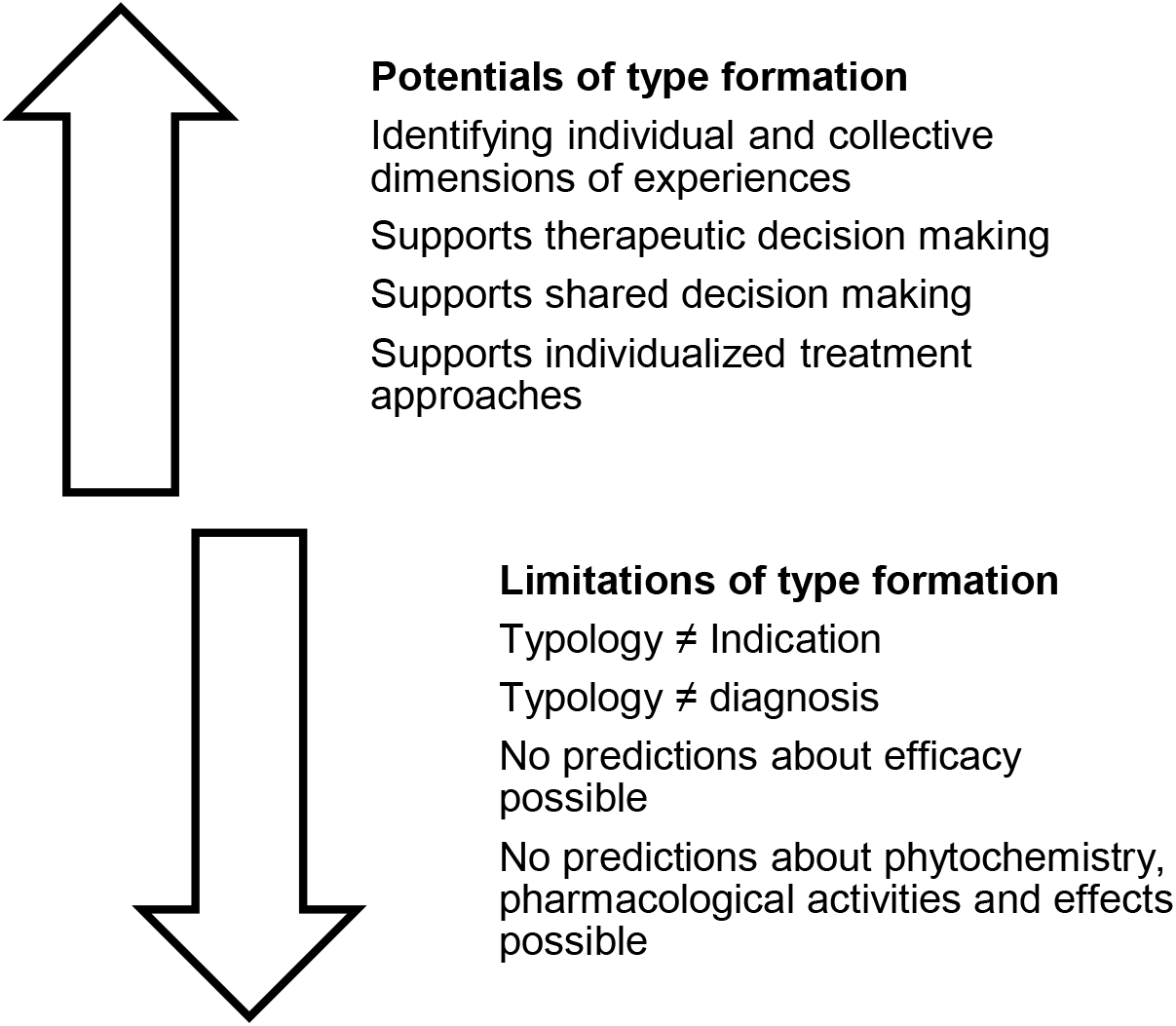
Potentials and limitations of type formation in the clinical context of phytomedicine and CIM in general

Type descriptions in phytotherapy and CIM neither equal indications for the herbal medicine preparations in question nor can they be a basis for diagnostics. Furthermore, no predictions about efficacy or pharmacological effects are possible based on type descriptions. Rather, we understand them as an additional tool supporting shared decision making and individualized treatment approaches in phytotherapy and CIM.

### Limitations and strengths of the two studies

From the 21 participants, twenty identified themselves as females and one as male. Although this is a limitation of the studies, it was somehow expectable for two reasons. First, it is well known that women seek more often consultation in CIM and are more willing to participate in respective scientific studies (see for example 22, 23). Second, based on clinical experience and traditional knowledge in phytotherapy (see for example 11), *Angelica archangelica* is known to be a medicinal herb especially suitable for women in transition and therefore mainly prescribed for women.

In both studies, we performed a purposeful sampling (14). However, it cannot be excluded that we might would have found more and different types or would have been able to differentiate subtypes from a bigger sample with more different characteristics.

We researched two specific ethanolic herbal extracts in a mainly hospital environment. Both certainly influenced our findings, so that future research should be conducted to corroborate the type descriptions with other ethanolic herbal preparations of *Passiflora incarnata* and *Angelica archangelica* in other health care settings with broader sample characteristics. Furthermore, it could be argued that participants probably were aware of the herbal medicine plant descriptions of the producer and reported their experiences accordingly. However, only three out of the 21 participants reported having knowledge of the producers’ herbal medicine plant descriptions.

Among the very few qualitative studies in the field of phytotherapy (e.g. 24, 25, 26), there are no other studies applying any form of type construction to our knowledge so far. Consequently, we contributed starting to fill this gap with our two studies about the use and everyday life experiences of two ethanolic herbal extracts, *Passiflora incarnata* and *Angelica archangelica*.

## Conclusion

Type construction might be an option to bridge qualitative health research results and therapeutic practice in complementary and integrative medicine by supporting processes of shared decision making and individualized treatment approaches in health settings.

According to our knowledge, having applied type construction to research data of everyday life experiences with the use of ethanolic herbal extracts is unique in the field of phytotherapy to date.

Future research should adopt a participatory implementation science approach to explore how type descriptions can be used and are experienced in therapeutic health care settings in complementary and integrative medicine.

## Supporting information

COREQ Checklist

## Data Availability

All data produced in the present study are available upon reasonable request to the authors

## List of abbreviations

CIM: complementary and integrative medicine

## Declarations

## Acknowledgments

We thank all the stakeholders for their participation in the two studies. In addition, we thank Dr. phil. Maja Dal Cero and Prof. Dr. med. Matthias Rostock for mentoring the studies.

## Conflicting Interests

The authors declare that they have no conflicting interests.

## Funding

The two studies were funded by Ceres Heilmittel AG, Switzerland. The funders had no role in study design, data collection and analysis, decision to publish, or preparation of the manuscript, but were regularly informed about the proceeding of the research.

## Ethical approval and consent to participate

All procedures performed in the two studies involving human participants were in accordance with the 1964 Helsinki declaration and its later amendments and with the ethical standards of the ethics committee of Zurich, Switzerland. Both studies did not fall under the regulation of the Human Research Act of Switzerland which was confirmed by the Ethics Committee of Zurich, Switzerland (Passiflora-Study: KEK-ZH-Nr. 59-2015, July 2015 / Angelica-Study: BASEC-Nr. Req-2017-00456, June 2017).

## Consent to participate and for publication

Written informed consent was obtained from all individual participants included in the study.

## Author Biographies

**Claudia Canella, MA** is a qualitative health researcher at the Institute for Complementary and Integrative Medicine of the University Hospital Zurich, Switzerland and a doctoral student at the Institute for Social Medicine, Epidemiology and Health Economics, Charité – Universitätsmedizin in Berlin, Germany.

**Dr. Balz Wolfensberger** is a senior lecturer and qualitative researcher at the Institute of Education, University of Zurich, Switzerland.

**Prof. Dr. med. Claudia Witt, MBA** is the clinical director and head of the Institute for Complementary and Integrative Medicine of the University Hospital Zurich, Switzerland and holds a full professorship at the University of Zurich, Switzerland.

## Author Contributions

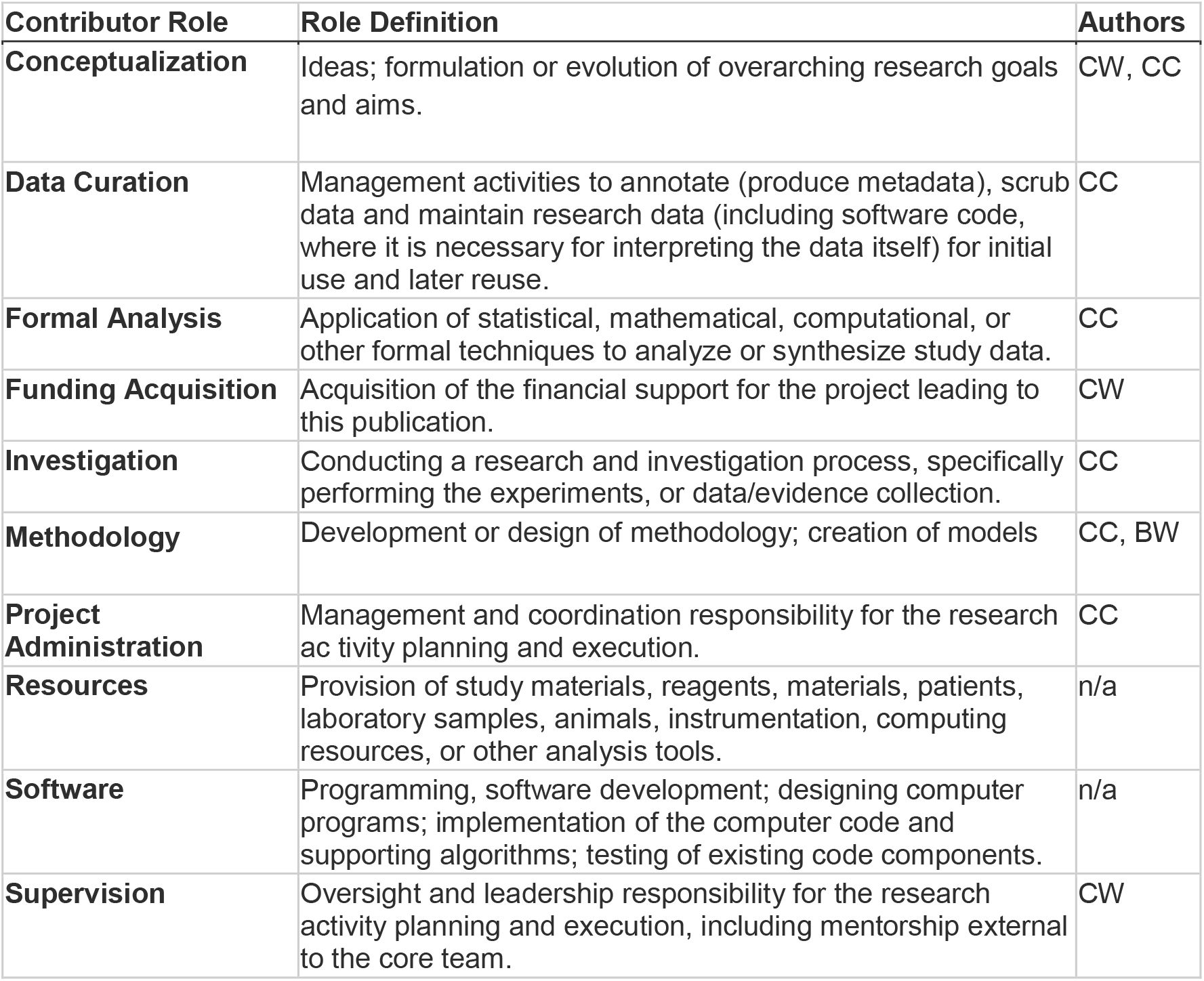
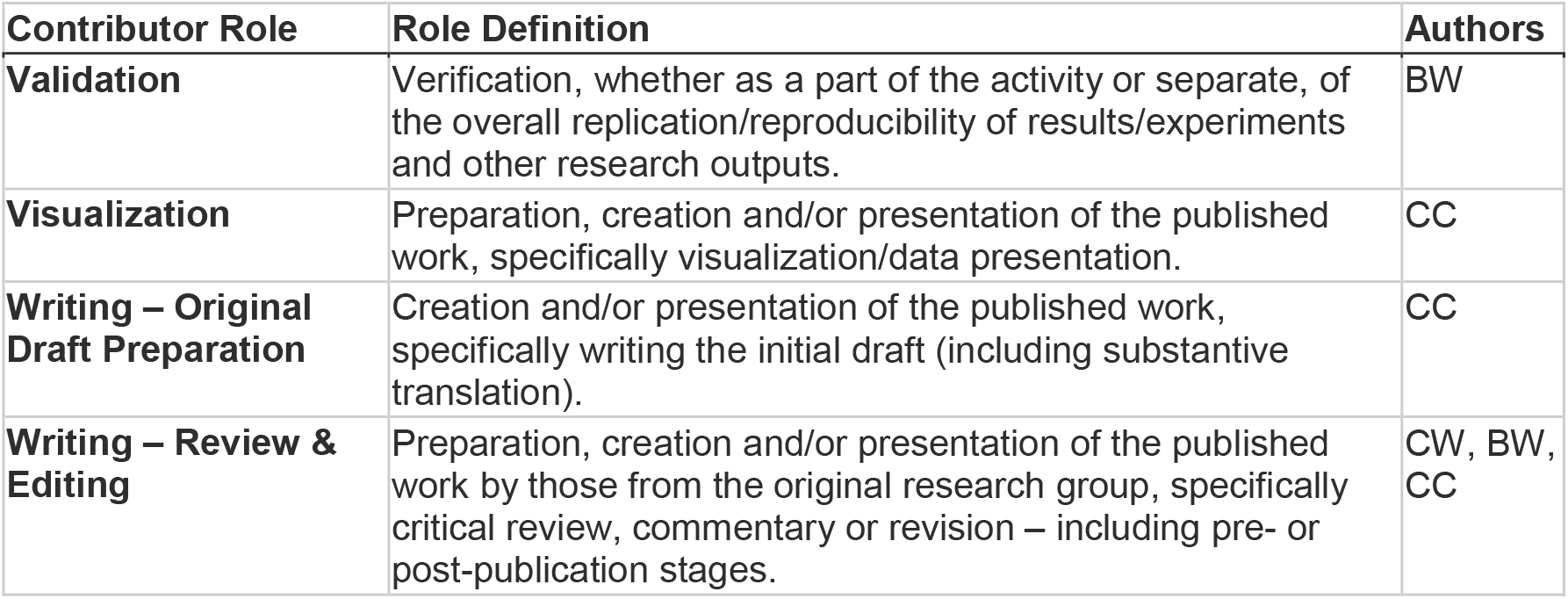

## Websites

https://www.cochrane.de/de/ebm, last accessed 03/19/21.

https://www.pcori.org/, last accessed 03/19/21.

## Notes

### Competing Interest Statement

The authors have declared no competing interest.

### Author Declarations

Ethical approval and consent to participate: All procedures performed in the two studies involving human participants were in accordance with the 1964 Helsinki declaration and its later amendments and with the ethical standards of the ethics committee of Zurich, Switzerland. Both studies did not fall under the regulation of the Human Research Act of Switzerland which was confirmed by the Ethics Committee of Zurich, Switzerland (Passiflora-Study: KEK-ZH-Nr. 59-2015, July 2015 / Angelica-Study: BASEC-Nr. Req-2017-00456, June 2017). Consent to participate and for publication: Written informed consent was obtained from all individual participants included in the study.

