## Supplementary material for "Type construction in qualitative health research and its relevance in complementary and integrative medicine. Two studies about patients’ experiences with herbal medicine preparations": COREQ Checklist

### Consolidated criteria for reporting qualitative studies (COREQ): 32-item checklist

| No | Item | Guide questions/description | Reported on |
|---|---|---|---|
| **Domain 1: Research team and reflexivity** | | | |
| Personal Characteristics | | | |
| 1. | Interviewer/facilitator | Which author/s conducted the interview or focus group? | Page Nr.: 6-7 Paragraphs: 120-156 |
| 2. | Credentials | What were the researcher's credentials? *E.g. PhD, MD* | See Paragraph “Author Biographies”, p.25-26 |
| 3. | Occupation | What was their occupation at the time of the study? | See Paragraph “Author Biographies”, p.25-26 |
| 4. | Gender | Was the researcher male or female? | See Paragraph “Author Biographies”, p.25-26 |
| 5. | Experience and training | What experience or training did the researcher have? | See Paragraph “Author Biographies”, p.25-26 |
| Relationship with participants | | | |
| 6. | Relationship established | Was a relationship established prior to study commencement? | Page Nr.: 6-7 Paragraphs: 120-156 |
| 7. | Participant knowledge of the interviewer | What did the participants know about the researcher? e.*g. personal goals, reasons for doing the research* | Page Nr.: 6-7 Paragraphs: 120-156 |
| 8. | Interviewer characteristics | What characteristics were reported about the interviewer/facilitator? e.g. *Bias, assumptions, reasons and interests in the research topic* | Page Nr.: 6-7 Paragraphs: 120-156 |
| **Domain 2: study design** | | | |
| Theoretical framework | | | |
| 9. | Methodological orientation and Theory | What methodological orientation was stated to underpin the study? *e.g. grounded theory, discourse analysis, ethnography, phenomenology, content analysis* | Page Nr.: 3-4 Paragraphs: 44-75 |

| No | Item | Guide questions/description | Reported on |
|---|---|---|---|
| Participant selection | | | |
| 10. | Sampling | How were participants selected? *e.g. purposive, convenience, consecutive, snowball* | Page Nr.: 6-8 Paragraphs: 120-156 |
| 11. | Method of approach | How were participants approached? e.*g. face-to-face, telephone, mail, email* | Page Nr.: 6-8 Paragraphs: 120-156 |
| 12. | Sample size | How many participants were in the study? | Page Nr.: 9-10 Paragraphs: 186-202 |
| 13. | Non-participation | How many people refused to participate or dropped out? Reasons? | Page Nr.: 8 Paragraphs: 151-156 |
| Setting | | | |
| 14. | Setting of data collection | Where was the data collected? e.*g. home, clinic, workplace* | Page Nr.: 6-8 Paragraphs: 120-156 |
| 15. | Presence of non-participants | Was anyone else present besides the participants and researchers? | No |
| 16. | Description of sample | What are the important characteristics of the sample? *e.g. demographic data, date* | Page Nr.: 9-10 Paragraphs: 186-202 |
| Data collection | | | |
| 17. | Interview guide | Were questions, prompts, guides provided by the authors? Was it pilot tested? | Page Nr.: 6-8 Paragraphs: 120-156 |
| 18. | Repeat interviews | Were repeat interviews carried out? If yes, how many? | Page Nr.: 6-8 Paragraphs: 120-156 |
| 19. | Audio/visual recording | Did the research use audio or visual recording to collect the data? | Page Nr.: 7 Paragraphs: 140 |
| 20. | Field notes | Were field notes made during and/or after the interview or focus group? | Yes |
| 21. | Duration | What was the duration of the interviews or focus group? | Page Nr.: 7 Paragraphs: 140 |
| 22. | Data saturation | Was data saturation discussed? | Page Nr.: 8 Paragraphs: 148-151 |
| 23. | Transcripts returned | Were transcripts returned to participants for comment and/or correction? | No |
| **Domain 3: analysis and findings**z | | | |
| Data analysis | | | |
| 24. | Number of data coders | How many data coders coded the data? | Page Nr.: 8-9 Paragraphs: 157-185 |
| 25. | Description of the coding tree | Did authors provide a description of the coding tree? | Page Nr.: 11-12 Paragraphs: 212-214 |

| No | Item | Guide questions/description | Reported on |
|---|---|---|---|
| 26. | Derivation of themes | Were themes identified in advance or derived from the data? | Page Nr.: 10-11 Paragraphs: 203-211 |
| 27. | Software | What software, if applicable, was used to manage the data? | Page Nr.: 8 Paragraphs: 158-159 |
| 28. | Participant checking | Did participants provide feedback on the findings? | Page Nr.: 9 Paragraphs: 184-185 |
| Reporting | | | |
| 29. | Quotations presented | Were participant quotations presented to illustrate the themes / findings? Was each quotation identified? *e.g. participant number* | Page Nr.: 10-17 Paragraphs: 203-327 |
| 30. | Data and findings consistent | Was there consistency between the data presented and the findings? | Page Nr.: 10-18 Paragraphs: 203-342 |
| 31. | Clarity of major themes | Were major themes clearly presented in the findings? | Page Nr.: 10-17 Paragraphs: 203-342 |
| 32. | Clarity of minor themes | Is there a description of diverse cases or discussion of minor themes? | Page Nr.: 10-17 Paragraphs: 203-342 |

Allison Tong, Peter Sainsbury, Jonathan Craig, Consolidated criteria for reporting qualitative research (COREQ): a 32-item checklist for interviews and focus groups, *International Journal for Quality in Health Care*, Volume 19, Issue 6, December 2007, Pages 349–357, https://doi.org/10.1093/intqhc/mzm042
